# Exact calculation of end-of-outbreak probabilities using contact tracing data

**DOI:** 10.1101/2023.08.31.23294914

**Authors:** NV Bradbury, WS Hart, FA Lovell-Read, JA Polonsky, RN Thompson

## Abstract

A key challenge for public health policy makers is determining when an infectious disease outbreak has finished. Following a period without cases, an estimate of the probability that no further cases will occur in future (the end-of-outbreak probability) can be used to inform whether or not to declare an outbreak over. An existing quantitative approach, based on a branching process transmission model, allows the end-of-outbreak probability to be approximated from disease incidence time series, the offspring distribution and the serial interval of the pathogen (the Nishiura method). Here, we show how the end-of-outbreak probability under the same transmission model can be calculated exactly if data describing who-infected-whom (the outbreak transmission tree) are available alongside the disease incidence time series. When such data are available, for example from contact tracing studies, our novel approach (the traced transmission method) is straightforward to use. We demonstrate this by applying the traced transmission method to data from previous outbreaks of Ebola virus disease and Nipah virus infection. For both outbreak datasets considered, we find that the traced transmission method would have determined that the outbreak was over more quickly than the Nishiura method. This highlights that consideration of contact tracing data may allow stringent control interventions to be relaxed quickly at the end of an outbreak, with only a limited risk of outbreak resurgence.

## Introduction

Infectious disease outbreaks require coordinated public health responses that limit the impacts of disease while avoiding unnecessary interventions. After an outbreak is brought under control, an important consideration is when the outbreak can be declared over safely. An end-of-outbreak declaration allows public health measures to be relaxed, but such a declaration must only be made when there is a low risk of a resurgence in cases [1,2]. World Health Organization (WHO) guidance for diseases such as Ebola virus disease (EVD) recommends that the acute phase of an outbreak is declared over when no new cases have been detected over a period of time that is equal to twice the theoretical maximum incubation period following the recovery or death of the last reported case (42 days for EVD [3]).

Simple rules for determining when to declare an outbreak over, based on fixed time periods without new cases, are straightforward to apply. However, the risk of a resurgence in cases in fact depends on specific features of the particular outbreak under consideration. Previous analyses have found that this risk depends on factors including the reproduction number, the extent of case underreporting, and the time between symptom onset and removal of the last detected case [4,5]. This indicates that there is a need for quantitative approaches that can be applied to guide decisions about when to declare an outbreak over, accounting for features of the outbreak under consideration.

There has been recent interest in using mathematical modelling to estimate the probability that no further cases of disease will occur in future (the *end-of-outbreak probability*), based on the observed outbreak data up to the current date [1,2]. If the end-of-outbreak probability can be estimated in real-time during an outbreak, then this quantity facilitates evidence-based removal of public health interventions. For example, an outbreak could be declared over as soon as the estimated end-of-outbreak probability exceeds a pre-specified threshold that is set based on the policy maker’s level of risk tolerance [1]. Several methods exist for estimating the end-of-outbreak probability [1,4–14]. The most commonly used approach [6–9], and therefore the basis from which we began our research here, was introduced by Nishiura *et al.* [6] (the *Nishiura method*) and is based on a branching process transmission model. The Nishiura method has the advantage of enabling the end-of-outbreak probability to be approximated straightforwardly using three inputs (Figure 1): (1) disease incidence time series (the number of cases recorded on each day of the outbreak up to the current time); (2) the serial interval distribution (the probability distribution describing the number of days between the symptom onset dates of an infector-infectee transmission pair); and (3) the offspring distribution (the probability distribution characterising the number of secondary cases generated by an infected host).

**Figure 1.**
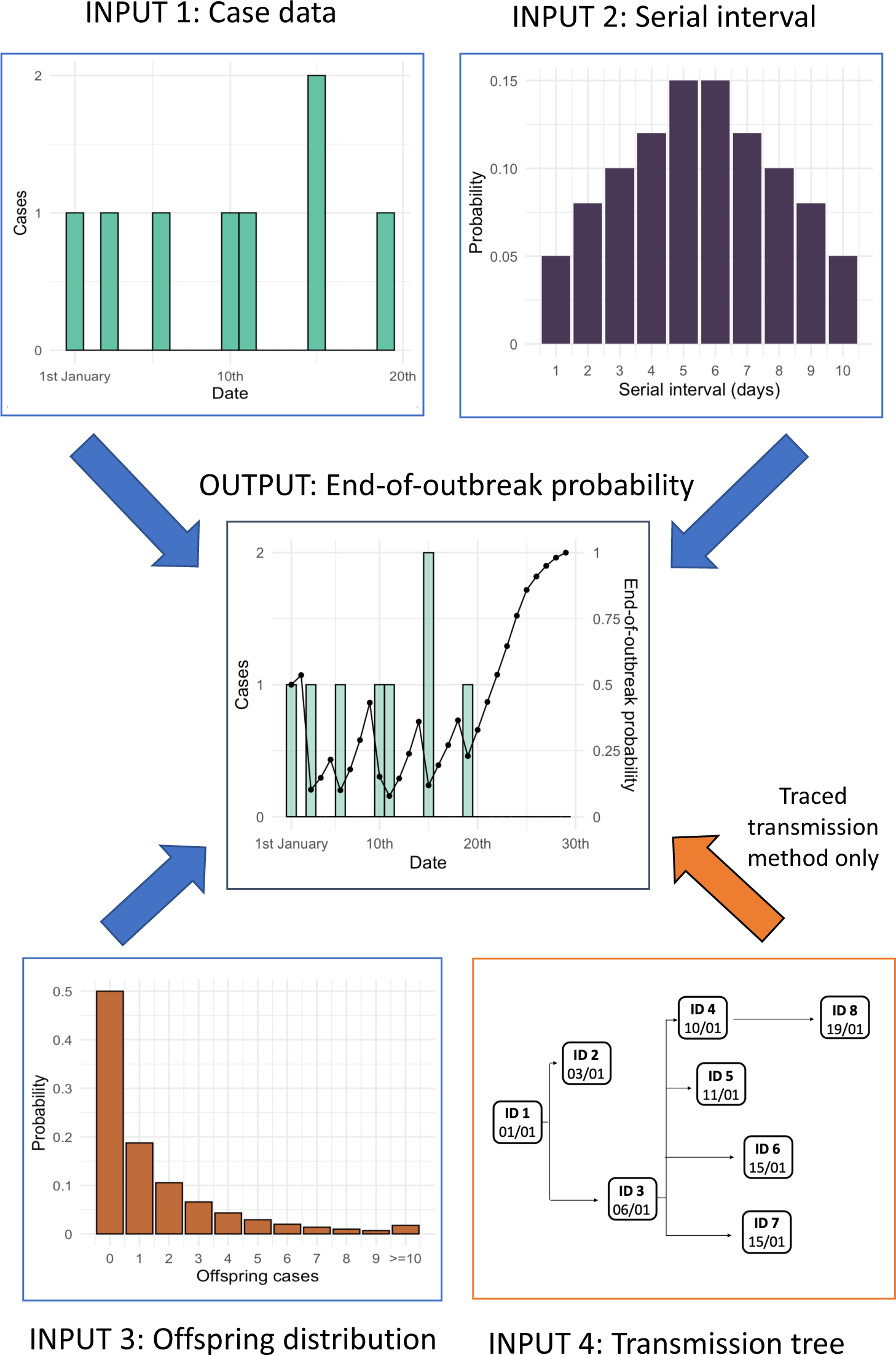
Schematic showing the inputs required for the Nishiura method and the traced transmission method for estimating the end-of-outbreak probability, and the type of output produced. Both methods require disease incidence time series (input 1), the serial interval distribution (input 2) and the offspring distribution (input 3). The traced transmission method also requires the outbreak transmission tree (input 4). Blue arrows therefore indicate the inputs required for both methods, and the orange arrow indicates the additional input required for the traced transmission method. The output of both methods is an estimate of the end-of-outbreak probability on a particular day (i.e., the probability that no further cases occur in future, based on the disease incidence time series data observed up to and including that day).

However, even if these inputs are known exactly, and transmission does indeed occur according to a branching process, the Nishiura method only provides an approximation of the end-of-outbreak probability (see Methods). Here, we provide a new approach (the *traced transmission method*) for calculating the end-of-outbreak probability exactly under the branching process transmission model used by Nishiura *et al*. [6] (Figure 1), which can be applied in scenarios in which information is available about who-infected-whom. Specifically, our approach uses the outbreak transmission tree, which can be obtained or estimated via contact tracing [7,15], in combination with the inputs to the Nishiura method. We consider two case studies of outbreaks of viral, zoonotic diseases: an EVD outbreak in Likati Health Zone, Democratic Republic of the Congo (DRC) in 2017 [16] and an outbreak of Nipah virus infection in Bangladesh in 2004 [17]. For each outbreak, we compare estimates of the end-of-outbreak probability obtained using the Nishiura method to analogous estimates using the traced transmission method. We demonstrate that our exact approach, with contact tracing data incorporated, leads to different estimates than the Nishiura method while remaining straightforward to apply. To encourage uptake of the novel traced transmission method to inform when outbreaks of a range of directly transmitted pathogens can be declared over, we also implement it in an online software application, available via https://github.com/nabury/End_of_outbreak_app.

## Methods

### Notation

Here, we define the notation used throughout this section and the remainder of this article:

- *p*(*y*) is the probability mass function of the offspring distribution (in other words, the probability that a randomly chosen infected individual infects *y* other people). In each outbreak case study, we assumed a negative binomial offspring distribution with mean *R* (the reproduction number) and dispersion parameter *k*, so that

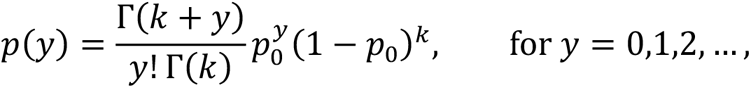
- where *p*_0_ = *R*/(*R* + *k*) and Γ(*z*) is the Gamma function. The choice of a negative binomial offspring distribution enables the effect of superspreading to be accounted for [18]. Specific parameter values used for the two case studies are given below.
- *w*(*x*) is the probability mass function of the (discrete) serial interval distribution (i.e., the probability that the interval between the symptom onset dates of an infector-infectee transmission pair is *x* days, where we assumed that only non-negative serial intervals can occur), and *F*(*x*) is the corresponding cumulative distribution function. For each outbreak case study, a discrete serial interval distribution was obtained by using the method described by Cori *et al.* [19] to discretise a published estimate of the continuous serial interval distribution (the distribution of time periods between the precise symptom onset times of infector-infectee transmission pairs). For further details, see web appendix 11 of that article.
- *t* is the current time, at which we want to estimate the end-of-outbreak probability (the probability that no cases occur after day *t*).
- The cases recorded up to and including the current time, *t*, are labelled with integer IDs *i* = 1,2, …, *m* (ordered by symptom onset date). The corresponding symptom onset dates are denoted by *t*_1_, *t*_2_, …, *t_m_*.
- *a_i_* is the number of recorded cases who were infected by individual *i* up to (and including) the current time, *t*.

### The end-of-outbreak probability

Below, we describe the Nishiura method and the traced transmission method for estimating the end-of-outbreak probability. Both of these methods are based on a branching process transmission model in which each infected host generates a number of cases that is sampled from the offspring distribution. These secondary cases then arise in the disease incidence time series after time periods (following the infector) that are sampled independently from the serial interval distribution. However, whereas the existing Nishiura method only approximates the end-of-outbreak probability under this transmission model (as explained below), the novel traced transmission method enables the end-of-outbreak probability to be calculated exactly whenever the outbreak transmission tree is known.

#### Nishiura method

Using the notation described in the previous subsection, in the Nishiura method [6] each case to date, *i*, is considered in turn. The offspring and serial interval distributions are used to calculate the probability, *q_i_*, that every secondary case generated by individual *i* develops symptoms no later than the current time, *t*, assuming nothing is known about the number (or symptom onset dates) of secondary cases already generated by individual *i* (up to time *t*).

Specifically,

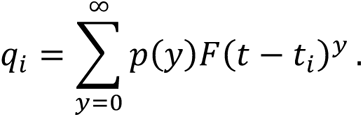

The end-of-outbreak probability can then be approximated by

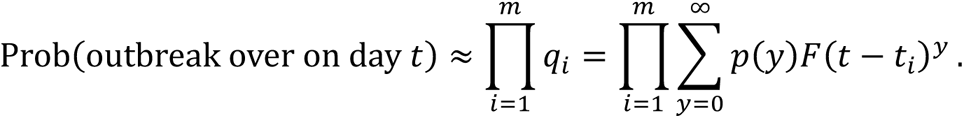

If the offspring distribution is a negative binomial distribution, then this formula simplifies to

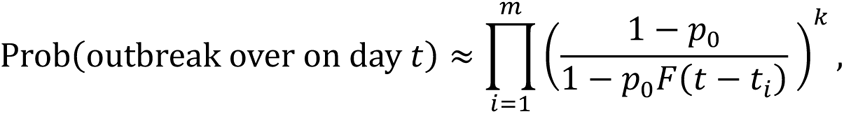

as shown in the Supplementary Material.

However, this is only an approximation of the end-of-outbreak probability under the assumed branching process transmission model because the remaining case data are not accounted for when calculating *q*_*i*_ (the probability that previous case *i* generates no future secondary cases). Even if the transmission tree is not known, the symptom onset dates of other recorded cases still provide some information about how many secondary cases may have been generated by each case *i*; this information is not used in the Nishiura method. In other words, the risk that a past case generates future infections is assumed to be independent of the number of infections that the individual has already generated, but this assumption may not hold in the branching process transmission model on which the Nishiura method is based. Specifically, under a negative binomial offspring distribution, the probability of an individual generating future cases increases with the number of cases generated to date – this reflects, for example, that a more infectious individual is likely to have generated more secondary cases to date (and is also more likely to generate future cases) than a less infectious individual who developed symptoms on the same date [18]. Therefore, the Nishiura method only approximates the end-of-outbreak probability, irrespective of whether or not the transmission tree is known.

#### Traced transmission method

If the transmission tree up to the current time, *t*, is known (so that, in particular, the number of secondary cases, *a_i_*, generated to date by each existing case, *i*, is known), then assuming that transmissions occur according to a branching process as described above, the end-of-outbreak probability can be calculated exactly and is given by

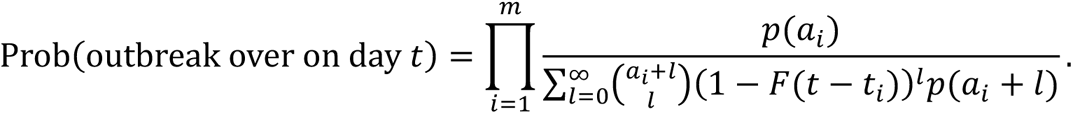

This expression is derived in the Supplementary Material. Again, in the case of a negative binomial offspring distribution, this can be simplified, here giving

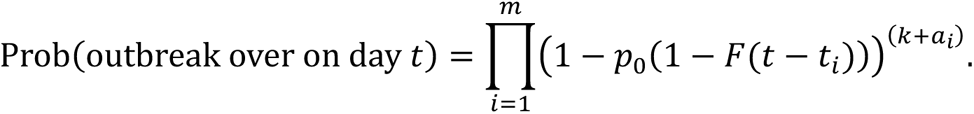

### Outbreak case studies

#### Case study 1: Ebola virus disease, Likati, Democratic Republic of the Congo

The first case study we considered is an EVD outbreak that occurred in the Likati Health Zone of DRC in 2017 [16]. Eight EVD cases were reported between 27 March and 11 May 2017, and symptom onset dates were recorded; five cases were confirmed and the remaining three were probable. Four of the infected individuals died [16]. The transmission tree was constructed using contact tracing and the symptom onset date of each case [16] (Figure 2A).

**Figure 2.**
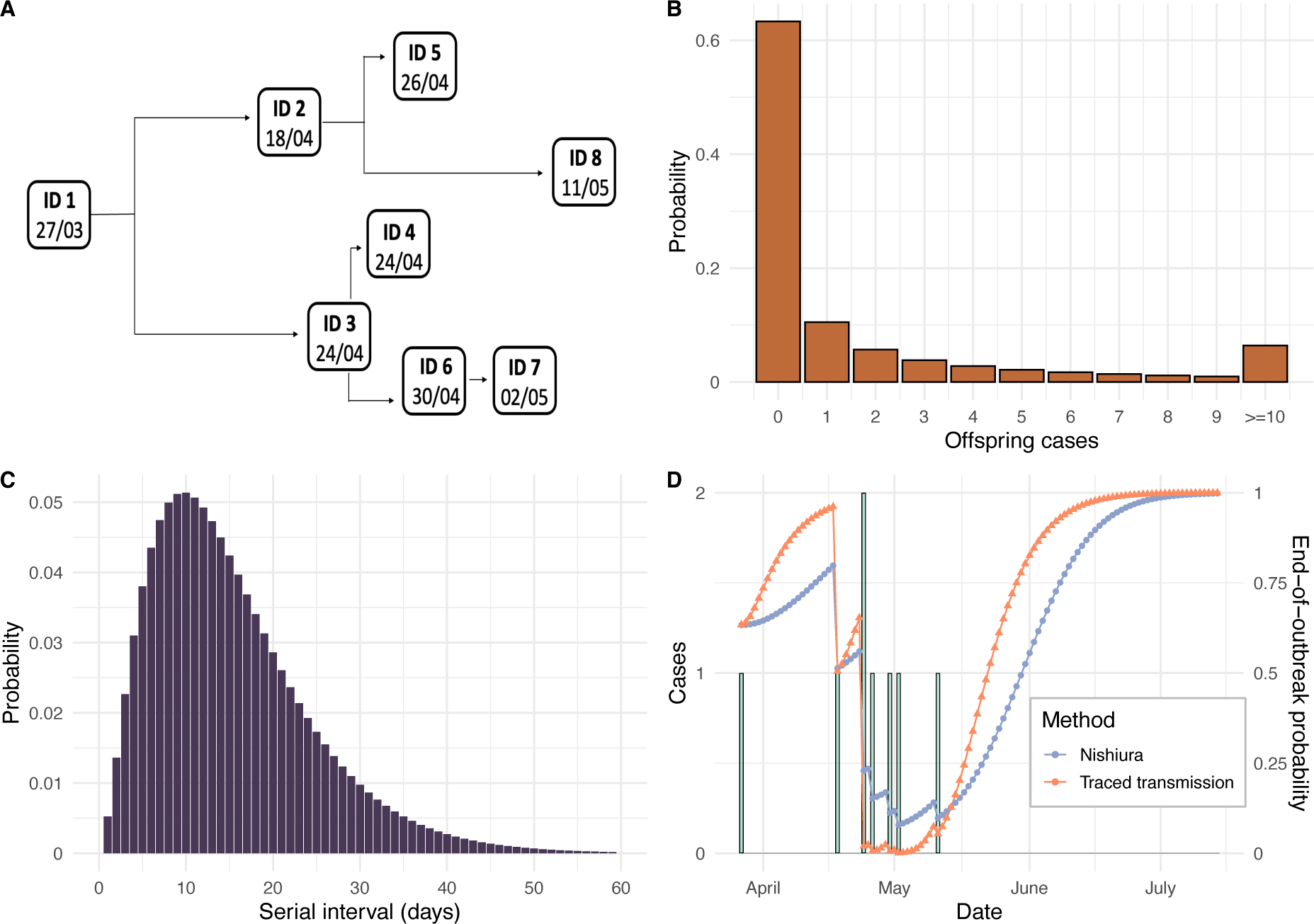
(A) Transmission tree for the 2017 EVD outbreak in the Likati Health Zone of DRC [16]. (B) Offspring distribution assumed for Ebola (negative binomial with reproduction number, *R* = 2.1 and dispersion parameter, *k* = 0.18 [20]). (C) Serial interval distribution assumed for Ebola. The continuous serial interval was assumed to be gamma-distributed with mean 15.3 days and standard deviation 9.3 days [20]. This distribution was then discretised using the method from [19]. (D) Estimated daily end-of-outbreak probabilities. Reported cases are represented by the green bars, with the left y-axis showing the daily number of cases. The line plots represent the estimated probability that the outbreak is over for each day of the outbreak for both the Nishiura method (blue) and the traced transmission method (orange). These probabilities are displayed on the right y-axis.

The Ebola offspring distribution was modelled as a negative binomial distribution with reproduction number *R* = 2.1 and dispersion parameter *k* = 0.18 [20] (Figure 2B). We assumed a gamma-distributed continuous serial interval distribution with mean 15.3 days and standard deviation 9.3 days [20], and discretised this distribution using the method of [19] as described above (Figure 2C).

End-of-outbreak probabilities were estimated each day (based on the data up to and including that day) from the symptom onset date of the first reported case until 100 days after the symptom onset date of the last reported case (total duration 146 days), using both the Nishiura method and the traced transmission method.

#### Case study 2: Nipah virus, Bangladesh

The second case study we considered is an outbreak of Nipah virus infection in the Faridpur district of Bangladesh between 19 February and 16 April 2004 [17]. Using laboratory testing and contact tracing, 36 cases were identified of which 23 were laboratory confirmed and 27 died [17]. The number of daily cases (by symptom onset date) peaked at nine on 1 April 2004 [17]. The probable transmission tree is shown in Figure 3A [17]. Two individuals (IDs *i* = 10 and *i* = 30) were not traced to any other cases [17], and were therefore considered as imported cases in our analyses (i.e., we assumed that they were not infected by any other case in the dataset). The individual with ID *i* = 6 was a local religious leader and had contact with 22 of the cases in this outbreak [17].

**Figure 3.**
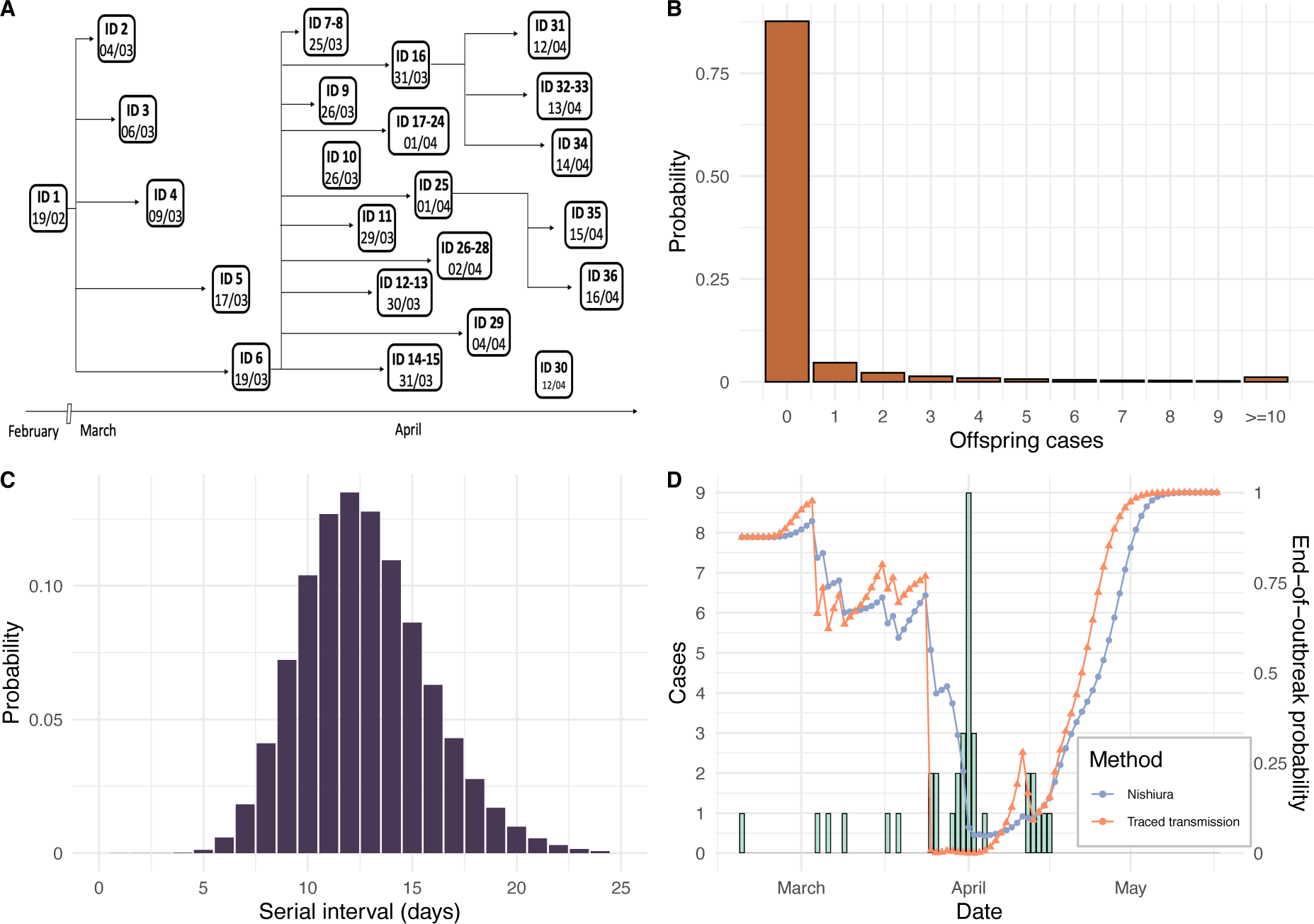
(A) Transmission tree for the 2004 outbreak of Nipah virus infection in Bangladesh. (B) Offspring distribution assumed for Nipah virus infection (negative binomial with reproduction number, *R* = 0.48 and dispersion parameter, *k* = 0.06 [21,22]). (C) Serial interval distribution assumed for Nipah virus infection. The continuous serial interval was assumed to follow a gamma distribution with mean 12.7 days and standard deviation 3.0 days [23]. This distribution was then discretised using the method from [19]. (D) Estimated daily end-of-outbreak probabilities. Reported cases are represented by the green bars, with the left y-axis showing the daily number of cases. The line plots represent the estimated probability that the outbreak is over for each day of the outbreak for both the Nishiura method (blue) and the traced transmission method (orange). These probabilities are displayed on the right y-axis.

The offspring distribution for this outbreak was modelled using a negative binomial distribution with reproduction number *R* = 0.48 and dispersion parameter *k* = 0.06 [21,22] (Figure 3B). We assumed a gamma distributed continuous serial interval distribution with mean 12.7 days and standard deviation 3.0 days [23], again discretised using the method of [19] (Figure 3C). End-of-outbreak probabilities were calculated daily from the date of the first case until 20 days after the last case (total duration 78 days).

In both case studies, we assumed that cases were reported on their symptom onset dates, thereby neglecting reporting delays when we estimated the end-of-outbreak probabilities. The symptom onset dates of reported cases are referred to as “case dates” in the remainder of this article.

## Results

### Real-time estimation of the end-of-outbreak probability

We first used both the Nishiura method and the novel traced transmission method to obtain end-of-outbreak probability estimates for the 2017 Likati EVD outbreak (Figure 2D). As would be expected, for both methods, the estimated end-of-outbreak probability increased over successive days without cases and decreased when new cases occurred.

While the general temporal trends were similar under both approaches, the extent of temporal variations in end-of-outbreak probability estimates was generally more pronounced for the traced transmission method. For example, on 17 April 2017 there had only been one reported case of EVD, with symptom onset date 21 days previously. The end-of-outbreak probability was estimated to be 0.8 using the Nishiura method, compared to a higher value of 0.96 using the traced transmission approach. On 2 May 2017, following six further cases, the estimated end-of-outbreak probability reached its lowest value for both approaches: 0.08 for the Nishiura method, and a lower value of 0.001 for the traced transmission method. Similarly, the end-of-outbreak probability increased more rapidly following the final case date for the traced transmission method than for the Nishiura method, with the probability first exceeding 0.99 on 22 June 2017 and 3 July 2017 using the two methods, respectively.

We then applied the two methods for estimating the end-of-outbreak probability to the data from the 2004 outbreak of Nipah virus infection in Bangladesh (Figure 3D). Again, following a cluster of new cases, the estimated end-of-outbreak probability generally fell lower for the traced transmission method than for the Nishiura method – for example, on 1 April 2004 (when there were nine new symptomatic cases, the highest daily number during the outbreak), the end-of-outbreak probability was 0.0001 for the traced transmission method and 0.07 for the Nishiura method. The end-of-outbreak probability also increased more rapidly following the final recorded case for the traced transmission method than for the Nishiura method.

### End-of-outbreak declaration thresholds

In principle, an outbreak could be considered over whenever the estimated end-of-outbreak probability exceeds a pre-determined threshold. This threshold can be set according to the policy maker’s willingness to accept a risk of future cases occurring (a lower threshold corresponds to a faster end-of-outbreak declaration, but with a higher risk that future cases occur). In Figure 4, we present plots showing the dates on which the 2017 Likati EVD outbreak would have been considered over for a range of end-of-outbreak probability threshold values. Results are shown for both the traced transmission method and the Nishiura method. Equivalent results for the 2004 outbreak of Nipah virus infection in Bangladesh are shown in Figure S1.

For each threshold considered, the two outbreaks would have been declared over earlier following the final case date using the traced transmission method than using the Nishiura method. Both methods suggested the EVD outbreak could potentially have been declared over earlier than the actual end-of-outbreak declaration date of 2 July 2017 [24] (42 days after the final case recovered, indicated as a green horizontal dash-dotted line in Figure 4) – the end-of-outbreak probability on 2 July 2017 was 0.998 for the traced transmission method and 0.989 for the Nishiura method.

**Figure 4.**
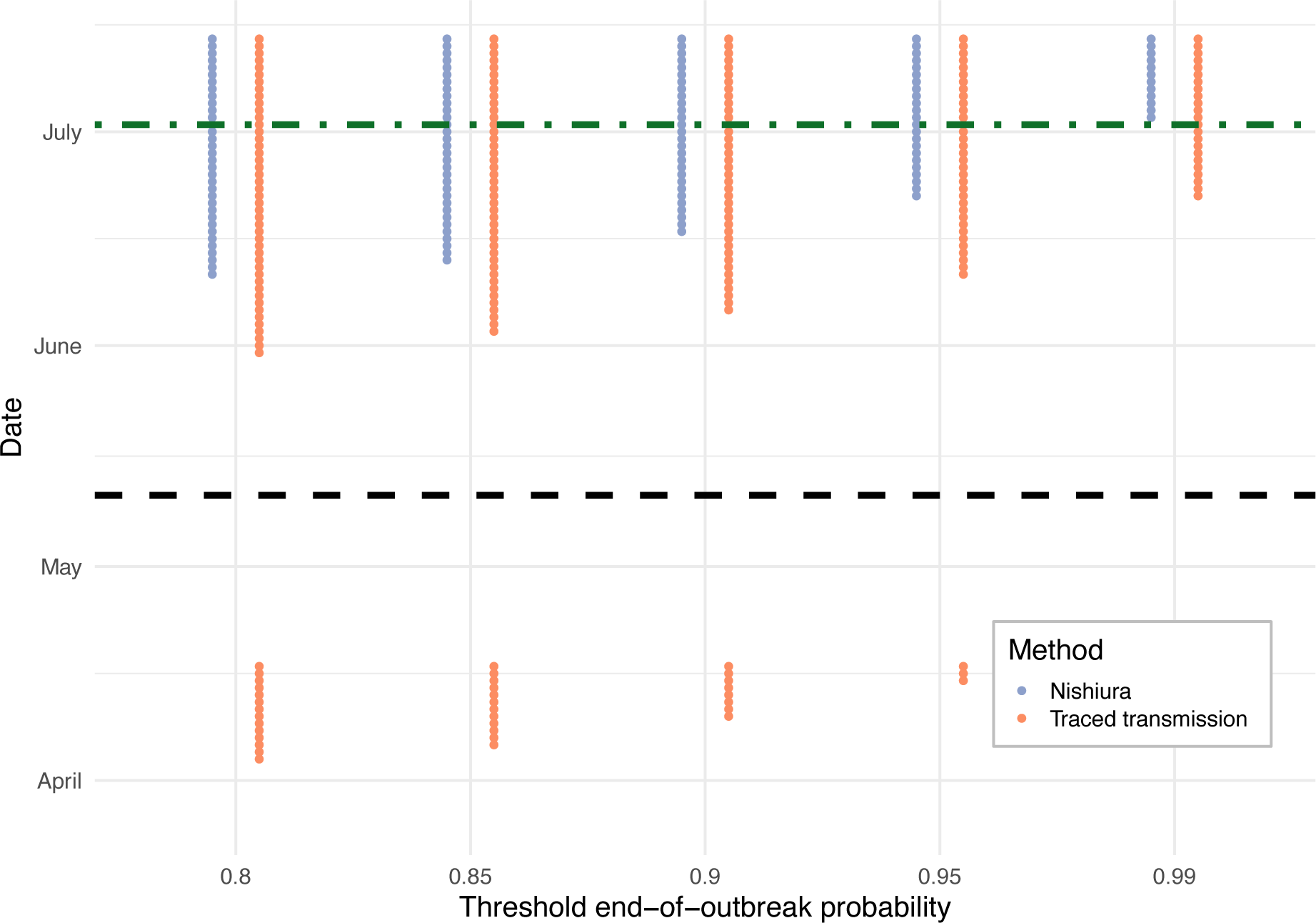
End-of-outbreak probability thresholds for the 2017 Likati EVD outbreak. The x-axis represents a range of end-of-outbreak probability thresholds, and the y-axis shows the outbreak dates on which these thresholds were exceeded by the estimated end-of-outbreak probability, for both the Nishiura method (blue) and the traced transmission method (orange). The date of the final recorded case (11 May 2017) is indicated as a black dashed line, and the actual end-of-outbreak declaration date (2 July 2017) [24] as a green dash-dotted line.

We note that, if the traced transmission method was used with an end-of-outbreak probability threshold of 0.96 or below, then the outbreak would have been considered over early in the outbreak, in April 2017, after only one case had occurred. A similar phenomenon can be seen when both the Nishiura method and the traced transmission method are applied to data from the Nipah virus infection outbreak (Figure S1). The occurrence of further cases when the transmission models suggested this to be unlikely is discussed below (see Discussion).

### Sensitivity of findings to the offspring distribution

Estimates of the reproduction number, *R* (representing overall transmissibility), and dispersion parameter, *k* (where lower values of *k* correspond to more overdispersed transmission; in other words, a greater degree of superspreading [18]), vary between outbreaks, even of the same infectious disease [18,22]. For example, the offspring distribution may differ due to different strains of a virus or behavioural characteristics of the affected population. We therefore investigated the effect of the assumed values of *R* and *k* on estimates of the end-of-outbreak probability using the Nishiura method and the traced transmission method, for the EVD and Nipah case studies (Figures S2 and S3). For each outbreak, we considered values of *R* and *k* both lower and higher than our assumed baseline values.

For both outbreaks, the assumed value of *k* (i.e., the extent of superspreading) had a particularly pronounced effect on the difference between the end-of-outbreak probability estimates under the Nishiura and traced transmission methods. In general, this difference was greater for lower values of the dispersion parameter, *k* (Figures S2 and S3). Higher values of *R* appeared to lead to larger differences between the methods than lower values of *R*.

## Discussion

Quantitative approaches for estimating the probability that an infectious disease outbreak has ended help policy advisors determine when the outbreak should be declared over [1,2]. Accurate estimation of the end-of-outbreak probability enables resource-intensive surveillance and control measures to be relaxed or removed as quickly as possible while limiting the risk of additional cases occurring. Here, we have developed a new approach (the traced transmission method) for estimating the end-of-outbreak probability in scenarios in which contact tracing enables reconstruction of the outbreak transmission tree (up to the current date). Our method uses the same branching process transmission model as an existing approximate approach for estimating the end-of-outbreak probability [6] (the Nishiura method), but unlike the Nishiura method, the traced transmission method gives the exact end-of-outbreak probability under this transmission model.

While we used the Nishiura method as the basis for our research given its previous use to calculate end-of-outbreak probabilities during outbreaks of a range of diseases (including MERS, EVD and COVID-19 [6–9]), we note that other methods for estimating the end-of-outbreak probability exist [4,5,10–14,25]. These methods have accounted for factors such as unreported cases [4,5,11] and temporal variations in the reproduction number [4,11]. While these other methods can be complex, sometimes requiring large numbers of simulations of stochastic epidemiological models to be run [4,5,25], a benefit of the Nishiura method is its straightforward application. The traced transmission method is similarly easy-to-use, allowing the end-of-outbreak probability to be estimated using a simple formula. We have developed an interactive, web-based app to facilitate future use of our approach (available via https://github.com/nabury/End_of_outbreak_app).

To demonstrate our method, we considered outbreaks of the Ebola and Nipah viruses as case studies. Both these viruses are zoonotic pathogens that cause sporadic outbreaks in humans, with high case fatality rates of 25-90% [26] and 40-75%, respectively [27]. Outbreaks are typically met with stringent control measures aiming to break chains of human-to-human transmission as rapidly as possible [26–28]. The question of when an outbreak can be declared over so that costly interventions can be safely relaxed or removed is therefore particularly pertinent to these viruses. Furthermore, as was the case for the two specific outbreaks we considered, intense contact tracing provides an opportunity to reconstruct outbreak transmission trees.

We found that estimates of the end-of-outbreak probability can vary substantially between our novel traced transmission method and the existing Nishiura method (Figures 2D and 3D). Specifically, the traced transmission method exhibited larger temporal variations in estimates of the end-of-outbreak probability during each outbreak, with the probability typically reaching lower values following clusters of new cases but then increasing more rapidly over successive days without any cases. The traced transmission approach therefore indicated that the two case study outbreaks could have been declared over earlier than suggested by the Nishiura method. The difference between the methods increased when assuming a smaller dispersion parameter, *k*, corresponding to a greater degree of superspreading (Figures S2 and S3).

While the difference in end-of-outbreak probability estimates between the two methods may be partially attributable to the fact that the traced transmission method leverages more data than the Nishiura method, this is unlikely to explain the consistent trends described in the previous paragraph fully. We note that, for the traced transmission method with a negative binomial offspring distribution, the probability of a recorded case generating no future cases is smaller if that individual has generated more secondary cases to date. This reflects the fact that an individual who has already generated more secondary cases may be more infectious and/or may have more contacts with susceptible individuals, and therefore may be more likely to generate future cases (either through future transmissions or past transmissions to individuals yet to develop symptoms), compared an individual who has generated fewer secondary cases to date. This effect is enhanced for a more overdispersed offspring distribution, when there is more variation between infected individuals in the number of secondary cases generated. On the other hand, the Nishiura method neglects information provided by the case data about how many secondary infections each case to date may have already generated. Even when the transmission tree is not known, some information is available about possible numbers of secondary cases generated by each case to date through the disease incidence time series, and this information is not used in the Nishiura method.

Both methods for estimating the end-of-outbreak probability considered here suggest a high probability that the Likati EVD outbreak had ended by the actual date on which the outbreak was declared over (based on current WHO guidance that recommends waiting for 42 days following the recovery or safe burial of the last recorded case). In comparison, one previous study recommended that the current 42 day waiting time guideline needed to be extended to ensure a high probability of an EVD outbreak being over at the time of an end-of-outbreak declaration [4], while another study found the appropriate waiting time to depend on the level of surveillance [5]. This second finding is consistent with our result here that a shorter waiting time before declaring the Likati EVD outbreak over may have been sufficient, since intensive contact tracing was undertaken.

For both case studies considered, our traced transmission method gives a high end-of-outbreak probability estimate immediately before the second case occurred (0.96 for the Likati EVD outbreak and 0.98 for the Bangladesh Nipah outbreak), and a similar phenomenon can be seen using the Nishiura method for the Nipah outbreak. While the subsequent occurrences of further cases may have indeed been realisations of unlikely events, other explanations for this finding are possible. First, the assumed offspring and serial interval distributions may not have been correct for the specific outbreaks considered – for example, a higher value of the dispersion parameter would lead to lower end-of-outbreak probability estimates on the corresponding days (see Figures S2 and S3). Alternatively, long gaps between the first and second recorded cases may have resulted from unrecorded intermediate cases. This is particularly likely early in an outbreak when intensive surveillance may not yet be in place. Extension of the traced transmission method to account for unreported cases and reporting delays, considering the sensitivity of the surveillance system and the time to put enhanced surveillance in place, is a target for future exploration, particularly as pathogens such as the Ebola virus tend to emerge in locations with weak surveillance. In addition, other possible areas for future work include accounting for uncertainty and/or temporal changes in the offspring and serial interval distributions.

In general, careful consideration should be given to the choice of probability threshold for an end-of-outbreak declaration, to appropriately balance the risk of an incorrect declaration with the economic and social costs of maintaining stringent outbreak controls for longer than necessary. One possibility is to take a stepped approach in which an initial end-of-outbreak declaration is made but surveillance measures are not removed completely until the estimated end-of-outbreak probability reaches a second, higher, threshold. Current policy for EVD requires heightened surveillance to be maintained for at least six months following an initial end-of-outbreak declaration [3].

In summary, we have developed a new approach for calculating the end-of-outbreak probability that robustly accounts for recorded transmission data. Application of our method indicates that two past outbreaks could have been declared over earlier than suggested by an existing approximate method. We hope that our approach is useful for informing end-of-outbreak declarations in future infectious disease outbreaks. The results from this modelling framework should be used as one of a range of sources of evidence to support public health decision making.

## AUTHORS’ CONTRIBUTIONS

NVM – formal analysis, investigation, software, validation, writing – review and editing.

WSH – formal analysis, investigation, software, validation, writing – original draft, writing – review and editing.

FAL – formal analysis, writing – review and editing.

JAP – methodology, writing – review and editing.

RNT – conceptualization, methodology, formal analysis, project administration, supervision, writing – original draft, writing – review and editing.

## ACKNOWLEDGEMENTS

Thanks to members of the Zeeman Institute for Systems Biology and Infectious Disease Epidemiology Research at the University of Warwick for useful discussions about this research.

## DATA AVAILABILITY

An interactive, web-based Shiny app was developed to conduct end-of-outbreak probability calculations using the traced transmission approach. This application and underlying R code are available at https://github.com/nabury/End_of_outbreak_app. All coding and analysis was conducted in the R programming language (version 4.0.2).

