## Supplementary Material for "Exact calculation of end-of-outbreak probabilities using contact tracing data"

### Supplementary Text

#### Nishiura method with a negative binomial offspring distribution

As described in the main text, the Nishiura method involves the approximate formula for the end-of-outbreak probability,

$$\text{Prob(outbreak over on day } t) \approx \prod_{i=1}^{m} \sum_{y=0}^{\infty} p(y) F(t-t_i)^y \,.$$

The notation used here is defined in the Methods section of the main text. In the special case of a negative binomial offspring distribution, we then have

$$\text{Prob(outbreak over on day } t) \approx \prod_{i=1}^{m} \sum_{y=0}^{\infty} \frac{\Gamma(k+y)}{y!\,\Gamma(k)} p_0^y (1-p_0)^k F(t-t_i)^y$$

$$= \prod_{i=1}^{m} \left(\frac{1-p_0}{1-p_0 F(t-t_i)}\right)^k \sum_{y=0}^{\infty} \frac{\Gamma(k+y)}{y!\,\Gamma(k)} \big(p_0 F(t-t_i)\big)^y \big(1-p_0 F(t-t_i)\big)^k$$

$$= \prod_{i=1}^{m} \left( \frac{1 - p_0}{1 - p_0 F(t - t_i)} \right)^k ,$$

with the final equality following because $\frac{\Gamma(k+y)}{y!\Gamma(k)} \left(p_0 F(t - t_i)\right)^y (1 - F(t - t_i) p_0)^k$ is the probability mass function of a negative binomial distribution.

**Traced transmission method**

Here, we derive an equation for the end-of-outbreak probability when the transmission tree is known up to the current time, $t$, assuming that transmissions occur according to a branching process. In addition to the notation introduced in the Methods of the main text, we introduce two further definitions:

- For each $0 \le s \le t$, $z_i(s)$ is the number of cases generated by individual $i$ that developed symptoms on calendar day $s$. The notation $z_i$ denotes the entire sequence of daily numbers of cases generated by individual $i$ (to date).
- $Y_i$ is a random variable giving the total number of secondary cases ever generated by individual $i$, including those occurring after the current time, $t$.

We first note that the end-of-outbreak probability depends only on the timing, $t_i$, of each case to date and on the corresponding daily secondary case frequency sequences, $z_i$ (in particular, the exact identities of the individuals infected by $i$ do not affect the end-of-outbreak probability). The end-of-outbreak probability, conditional on the available data, is then

$$\text{Prob}(\text{ no cases after day } t \mid (m \text{ total cases up to and including time } t), t_1, \dots, t_m, z_1, \dots, z_m )$$

$$= \text{Prob}( Y_1 = a_1, \dots, Y_m = a_m \mid t_1, \dots, t_m, z_1, \dots, z_m )$$

$$= \frac{\text{Prob}( Y_1 = a_1, \dots, Y_m = a_m, z_1, \dots, z_m \mid t_1, \dots, t_m )}{\text{Prob}( z_1, \dots, z_m \mid t_1, \dots, t_m )}$$

$$= \prod_{i=1}^{m} \frac{\text{Prob}( Y_i = a_i, z_i \mid t_i )}{\text{Prob}( z_i \mid t_i )}$$

$$= \prod_{i=1}^{m} \frac{\text{Prob}(\,Y_i = a_i, z_i \mid t_i\,)}{\sum_{l=0}^{\infty} \text{Prob}(\,Y_i = a_i + l, z_i \mid t_i\,)},$$

where the third equality follows by the assumption that transmissions occur according to a branching process (so that, in particular, different infected individuals generate transmissions independently of each other).

Now, we have

$$\text{Prob}(\,Y_i = a_i + l, z_i \mid t_i\,)$$

$$= \text{Prob}(\,z_i \mid t_i, Y_i = a_i + l\,) \times \text{Prob}(\,Y_i = a_i + l \mid t_i\,)$$

$$= \binom{a_i + l}{z_i(0), z_i(1), \ldots, z_i(t), l} \left( \prod_{s=0}^{t} w(s - t_i)^{z_i(s)} \right) (1 - F(t - t_i))^l \times p(a_i + l).$$

In this expression, $w(s - t_i)^{z_i(s)}$ gives the probability that $z_i(s)$ specified secondary cases generated by individual $i$ occur on day $s$ (defining this term to equal 1 when both $w(s - t_i)$ and $z_i(s)$ are zero), $(1 - F(t - t_i))^l$ gives the probability that $l$ specified secondary cases occur after the current day, $t$, and the multinomial coefficient,

$$\binom{a_i + l}{z_i(0), z_i(1), \ldots, z_i(t), l} = \frac{(a_i + l)!}{z_i(0)!\, z_i(1)! \ldots z_i(t)!\, l!},$$

gives the number of ways in which the $(a_i + l)$ total infectees can be divided into cases occurring on each day up to day $t$ and those occurring after day $t$.

Substituting and simplifying, the end-of-outbreak probability is therefore

$$\text{Prob}(\text{ no cases after time } t \mid (m \text{ total cases up to and including time } t), t_1, \ldots, t_m, z_1, \ldots, z_m\,)$$

$$= \prod_{i=1}^{m} \frac{p(a_i)}{\sum_{l=0}^{\infty} \binom{a_i + l}{l} (1 - F(t - t_i))^l p(a_i + l)}.$$

Further simplification is again possible for a negative binomial offspring distribution. In that case, we have

$$\sum_{l=0}^{\infty} \binom{a_i + l}{l} (1 - F(t - t_i))^l p(a_i + l)\,,$$

$$= \sum_{l=0}^{\infty} \frac{(a_i + l)!}{l!\, a_i!} (1 - F(t - t_i))^l \frac{\Gamma(k + a_i + l)}{(a_i + l)!\,\Gamma(k)} p_0^{a_i + l} (1 - p_0)^k\,,$$

$$= \frac{\Gamma(k + a_i)}{a_i!\,\Gamma(k)} p_0^{a_i} (1 - p_0)^k \big(1 - p_0(1 - F(t - t_i))\big)^{-(k + a_i)},$$

$$\times \sum_{l=0}^{\infty} \frac{\Gamma\big((k + a_i) + l\big)}{l!\,\Gamma(k + a_i)} \big(p_0(1 - F(t - t_i))\big)^l \big(1 - p_0(1 - F(t - t_i))\big)^{(k + a_i)}\,,$$

$$= p(a_i)\big(1 - p_0(1 - F(t - t_i))\big)^{-(k + a_i)},$$

since the final summand is the probability mass function of a negative binomial distribution. Therefore,

$$\text{Prob( no cases after day } t \mid t_1, \dots, t_m, z_1, \dots, z_m \text{ )} = \prod_{i=1}^{m} \big(1 - p_0(1 - F(t - t_i))\big)^{(k + a_i)}.$$

### Supplementary Figures

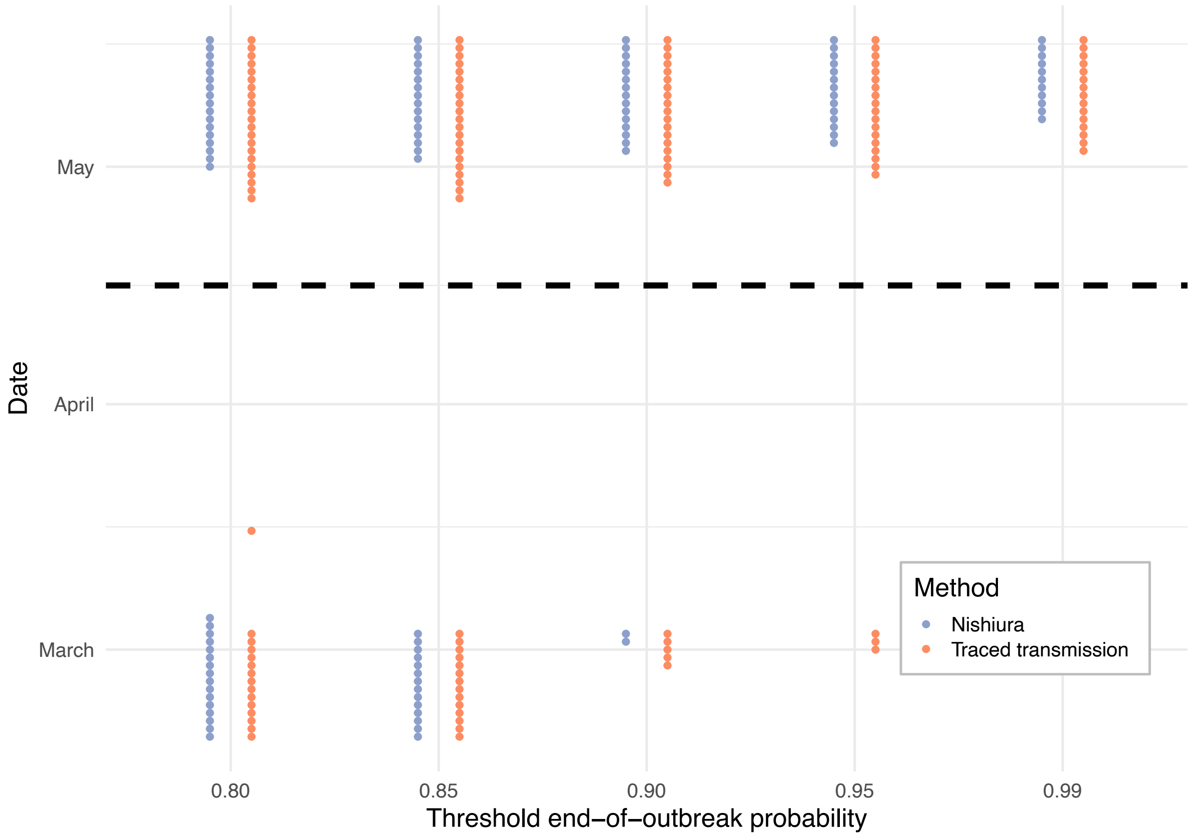


**Figure S1. End-of-outbreak probability thresholds for the 2004 Nipah virus infection outbreak in Bangladesh.** The x-axis represents a range of end-of-outbreak probability thresholds, and the y-axis shows the outbreak dates on which these thresholds were exceeded by the estimated end-of-outbreak probability, for both the Nishiura method (blue) and the traced transmission method (orange). The date of the final recorded case (16 April 2004) is indicated as a black dashed line.

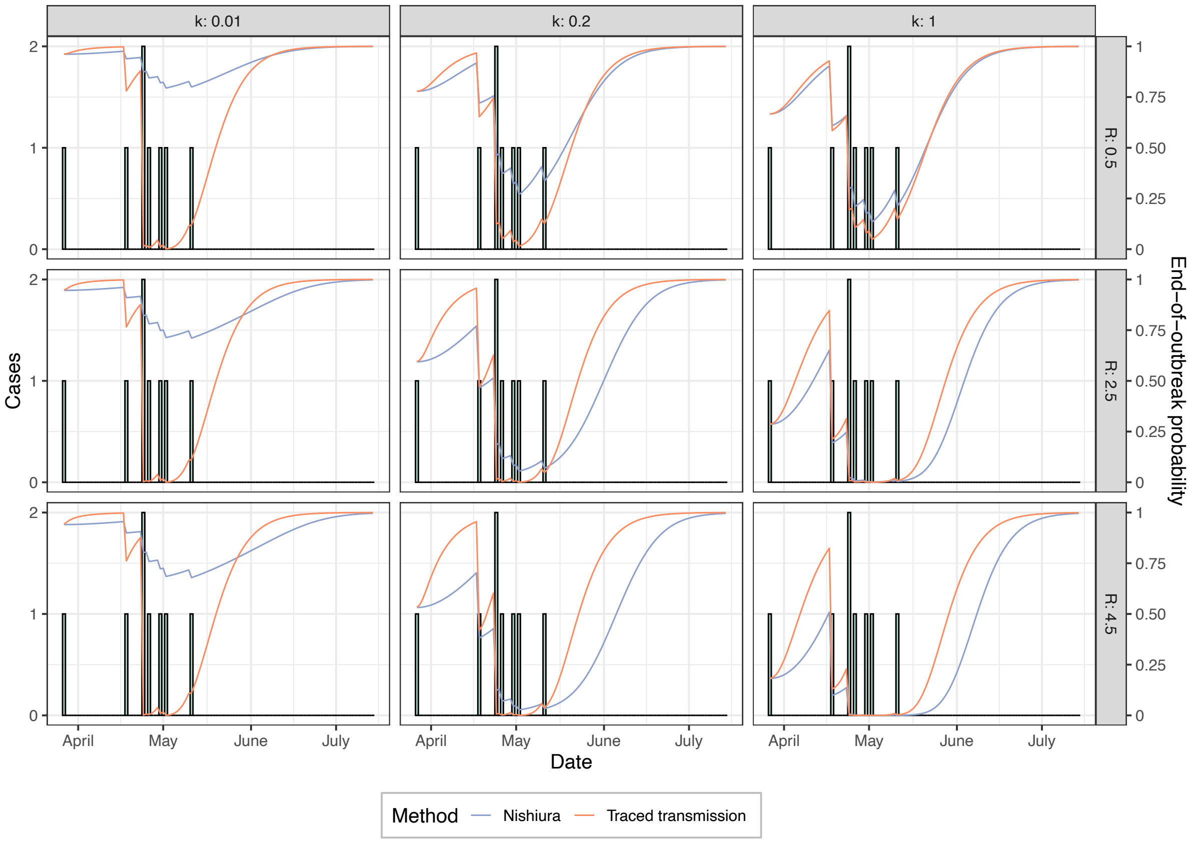


**Figure S2.** Daily end-of-outbreak probability estimates for the 2017 EVD outbreak in Likati, DRC, for different values of the reproduction number, $R$, and dispersion parameter, $k$, between panels. Rows represent values of $R$ from 0.5 (top) to 4.5 (bottom), and columns represent values of $k$ from 0.01 (left) to 1 (right).

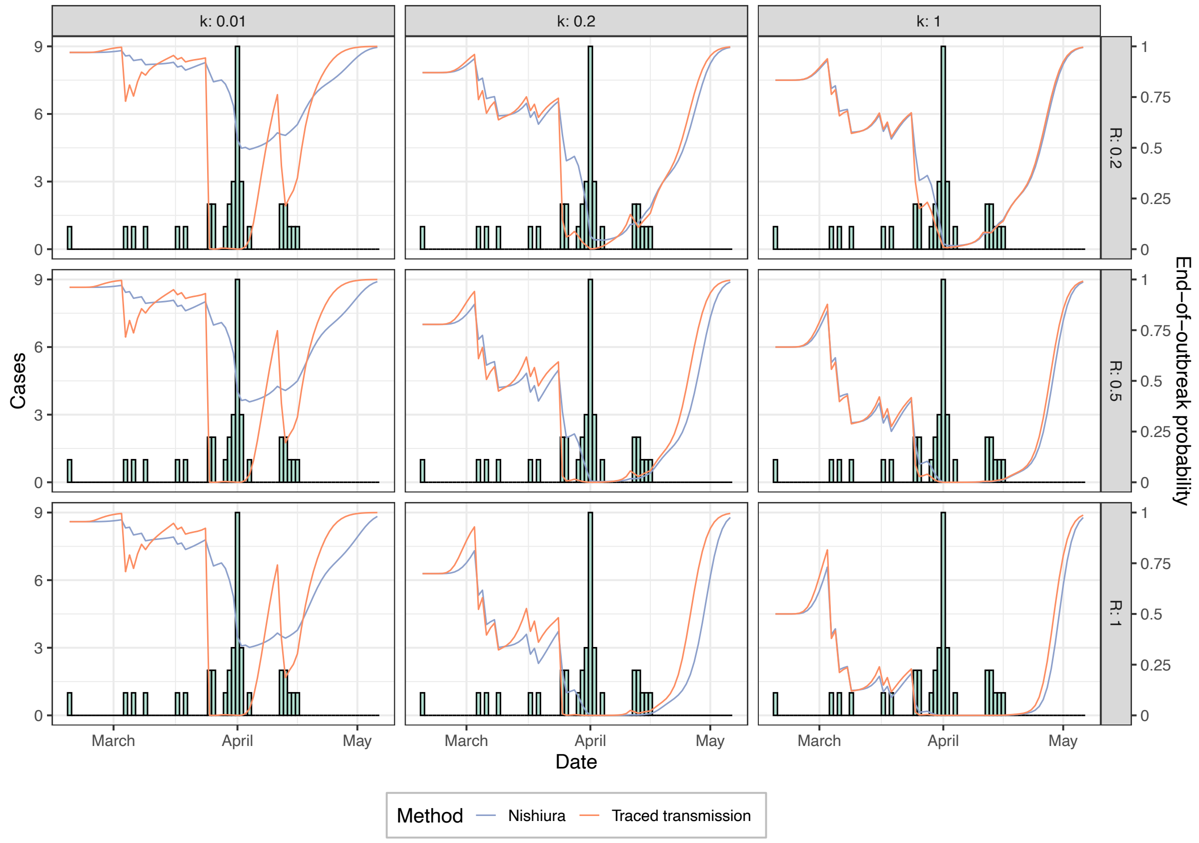


**Figure S3.** Daily end-of-outbreak probability estimates for the 2004 Nipah virus infection outbreak in Bangladesh, for different values of the reproduction number, $R$, and dispersion parameter, $k$, between panels. Rows represent values of $R$ from 0.2 (top) to 1 (bottom), and columns represent values of $k$ from 0.01 (left) to 1 (right).
